# The Development, Content Validity and Evaluation of an African-Contextual Back Pain Self-Management Model (AC-BP-SMM): *The Delphi Technique*

**DOI:** 10.64898/2026.03.09.26347978

**Authors:** Loveness A. Nkhata, Geoffrey Moyo, Olaitan Johnson Balogun, Margaret M. Mweshi

## Abstract

**Background:** Back pain is a leading occupational health problem among healthcare workers globally, with a disproportionately higher burden in low- and middle-income countries (LMICs). Existing theoretical models including the Fear-Avoidance Model, Biopsychosocial Model, and Patient Activation Theory originate from high-income country contexts and insufficiently represent the cultural, structural, and organisational realities of African healthcare settings.

**Aim:** To develop and evaluate the content validity of an African-Contextual Back Pain Self-Management Model (AC-BP-SMM) using a two-stage approach: a framework synthesis grounded in a 15-year programme of empirical research from Zambia, followed by content validity evaluation through the Delphi technique.

**Methods:** Stage 1 drew on 17 studies (N > 1,000 participants) conducted in Zambia between 2010 and 2025, comprising epidemiological surveys, qualitative research, psychometric validation, systematic reviews, and digital health intervention trials. The five analytical phases that guided model development were: data immersion and familiarisation, concept identification and preliminary framework development, systematic indexing of data, integration with existing theoretical frameworks, and model refinement. Stage 2 involved a two-round Delphi study with a purposively selected panel of 18 experts comprising physiotherapists, occupational health specialists, back pain researchers, and healthcare policy practitioners from 10 countries across Africa, Europe, and North America who rated each model component on relevance, clarity, and representativeness using a 4-point Likert scale. Content Validity Index (CVI) scores were calculated per item (I-CVI) and for the overall model (S-CVI/Ave); consensus was defined a priori as I-CVI ≥ 0.80. The framework synthesis identified five sequential model components — Cultural and Occupational Context, Back Pain Beliefs, Coping Strategies, Participant Activation, and Self-Management Behaviours — and a persistent knowledge-action gap (40–45 percentage points) where healthcare workers possess strong back-pain knowledge (78–85%) but cannot sustain self-management behaviours (35–42%). Four moderators (educational interventions, digital health tools, workplace culture, and system-level factors) influence pathway strength. The Delphi process achieved consensus across two rounds: the overall S-CVI/Ave was 0.93, with all 17 items achieving I-CVI ≥ 0.80 in the final round, and inter-rater agreement of κ = 0.82 (95% CI 0.74–0.90). The AC-BP-SMM is the first empirically grounded, content-valid conceptual model tailored for back pain self-management in African healthcare settings. By foregrounding context as a primary determinant and operationalising the knowledge-action gap, the model provides a validated theoretical blueprint for culturally appropriate multi-component interventions targeting both beliefs and structural barriers, with implications for occupational health policy, digital health intervention design, and future implementation research.

## 1. Background

Low back pain is among the most frequent occupational conditions globally, with lifetime prevalence frequently exceeding 60–80% among nurses and allied health professionals [1–9]. Healthcare workers face substantial physical and psychosocial demands, including manual patient lifting, awkward postures, prolonged standing, shift work, and high emotional load, all of which increase the risk of both acute and chronic back pain [1–4,10]. The consequences span reduced productivity, absenteeism, presenteeism, and premature workforce exit, with significant economic burden for health systems globally [9,11,12].

The burden is amplified in LMICs, where fragile health systems, severe staff shortages, and limited ergonomic infrastructure constrain prevention and management [13–17]. In Sub-Saharan Africa (SSA), studies consistently report high prevalence of low back pain among nurses, caregivers, and physiotherapy personnel [2,4,18–22]. Nurses in Zambia often manage 30–40 patients per shift while performing manual handling without assistive devices [17,19,23]. Long shifts, fatigue, and extensive domestic responsibilities intensify risk and reduce opportunities for self-management [9,24,25].

Across Zambia, Uganda, Kenya, South Africa, Nigeria, Pakistan, Brazil, and Ghana, a persistent knowledge-action gap has been documented: workers understand evidence-based management but rarely implement it because of contextual barriers [7,13–16,19–21,26–29]. Prominent theoretical frameworks such as the Fear-Avoidance Model [30], Biopsychosocial Model [27], and Patient Activation Model [28] were developed in high-income settings and make assumptions about individual autonomy, resource availability, and care access that do not reflect LMIC realities. They inadequately capture cultural norms prevalent across SSA stoicism, spiritual coping, occupational self-sacrifice, and collectivist responsibility that shape pain interpretations and behaviour [3,14,15,29,31,32]. There is, therefore, a critical need for an empirically grounded, African-contextual model with established content validity. This study reports the development and content validity evaluation of the African-Contextual Back Pain Self-Management Model (AC-BP-SMM) using a two-stage approach: framework synthesis followed by the Delphi technique.

### 1.1 Rationale for an African-Contextual Model

Western models assume substantial individual autonomy, the ability to control daily routines, schedules, and healthcare decisions. In LMICs, healthcare workers face rigid institutional demands, cultural expectations of endurance, and limited decision-making authority over working conditions. A nurse in Zambia cannot refuse patient assignments when in pain, nor can an agricultural worker delay planting season because of back discomfort [11,14,15,18,33]. Second, Western models assume resource availability, access to physiotherapy, ergonomic equipment, and occupational health services. In SSA, physiotherapy ratios may be as low as 1:500,000 compared to 1:1,500 in high-income countries [13,16]. Third, Western models prioritise individualistic orientations, personal beliefs, self-efficacy, and individual responsibility whereas, African contexts emphasise collectivist values, family obligations, and spiritual interpretations of suffering [3,14,15,24,31,32]. Fourth, and most critically, existing models assume that correcting misbeliefs or increasing knowledge leads to behaviour change, whereas LMIC evidence consistently demonstrates a knowledge-action gap driven by structural constraints rather than knowledge deficits [7,13–16,19–21,26–29]. The AC-BP-SMM was developed to address these limitations by foregrounding context as the primary determinant.

### 1.2 Purpose of the Model

The AC-BP-SMM serves four interconnected purposes: (1) to articulate the mechanisms through which cultural and structural context shapes back pain self-management behaviour in African healthcare settings (Explanatory); (2) to operationalise the knowledge-action gap as a measurable, structurally driven outcome distinct from knowledge deficits (Theoretical); (3) to provide a blueprint for designing multi-component interventions that simultaneously target beliefs and structural barriers (Practical); and (4) to produce testable hypotheses for future implementation and validation research across LMIC populations (Generative).

### 1.3 Generalisability beyond Healthcare Workers

While developed from healthcare worker data, the AC-BP-SMM’s core constructs — cultural beliefs, knowledge-action gaps, structural barriers, and moderating factors — are theoretically applicable to other occupational groups in LMICs. Agricultural workers face analogous constraints: they understand that proper lifting reduces injury risk but cannot implement this during planting season when speed determines productivity [25,32]. Urban manual labourers face knowledge-action gaps driven by piece-rate payment systems that penalise rest breaks, lack of labour protections, and informal employment without sick leave [34,35]. The underlying pattern persists across contexts: knowledge exists but structural constraints prevent its translation into sustained behaviour. This generalisability informs ongoing pilot research with rural agricultural populations in Zambia, with findings expected in 2026.

## 2. Methods

The study employed a two-stage mixed-methods design: Stage 1 used a framework synthesis approach for model development; Stage 2 used the Delphi technique for content validity evaluation.

### 2.1 Stage 1: Model Development — Framework Synthesis

#### 2.1.1 Study Design and Methodological Framework

A framework synthesis approach [10,36] was employed to develop a comprehensive conceptual model explaining back pain self-management among healthcare workers in African LMIC contexts. Framework synthesis is suited for explanatory theory-building from diverse evidence sources [37], allowing iterative theory development through integration of qualitative, quantitative, and intervention evidence [36]. This approach was selected because the research question required explanatory understanding rather than effect estimation; the evidence base spanned multiple methodologies requiring integration rather than aggregation; and a 15-year longitudinal research programme provided cumulative insights.

### 2.1.2 Data Sources

The model is grounded in 17 studies conducted in Zambia between 2010 and 2025, involving more than 1,000 participants across diverse healthcare settings. Studies spanned cross-sectional epidemiological surveys, qualitative research, psychometric validation, systematic and scoping reviews, and intervention trials. Table 1 provides a comprehensive overview.

**Table 1:** Overview of Data Sources Informing Model Development (2010–2025)

| Category of Study | Description | Studies Included | Population / Setting |
| --- | --- | --- | --- |
| Cross-sectional epidemiological studies | Documented prevalence, correlates, and occupational determinants of back pain among healthcare workers. | [2,4,11,19,20] | Nurses and physiotherapy personnel in tertiary, secondary, and primary care facilities. (N = 563) |
| Qualitative studies | Explored lived experiences, cultural interpretations, occupational constraints, coping strategies, and belief systems. | [3,14,15] | Nurses, physiotherapists, and caregivers across multiple hospitals. (N = 87) |
| Psychometric studies | Validated back pain belief measures and adapted culturally appropriate frameworks for local use. | [5] | Nurses in tertiary and secondary facilities. (N = 185) |
| Systematic and scoping reviews | Evaluated evidence for self-management interventions and exercise-based strategies. | [16,18] | Published studies across global and SSA contexts. (systematic review design; primary N not applicable) |
| Intervention studies | Examined effects of educational campaigns and WhatsApp-based digital health on back pain beliefs, coping, and activation. | [6,13,26,33] | Nurses in Lusaka and other hospital settings. (N = 387) |
| **Total** | **Full body of work informing conceptual model development.** | **17 studies total** | **Over 1,000 participants — nurses, physiotherapists, caregivers, and allied health workers in Zambia.** |

#### 2.1.3 Framework Synthesis Procedure

The synthesis followed five systematic phases adapted from Carroll et al. [36] and operationalised for a longitudinal research programme (Fig 1).

**Fig 1.**
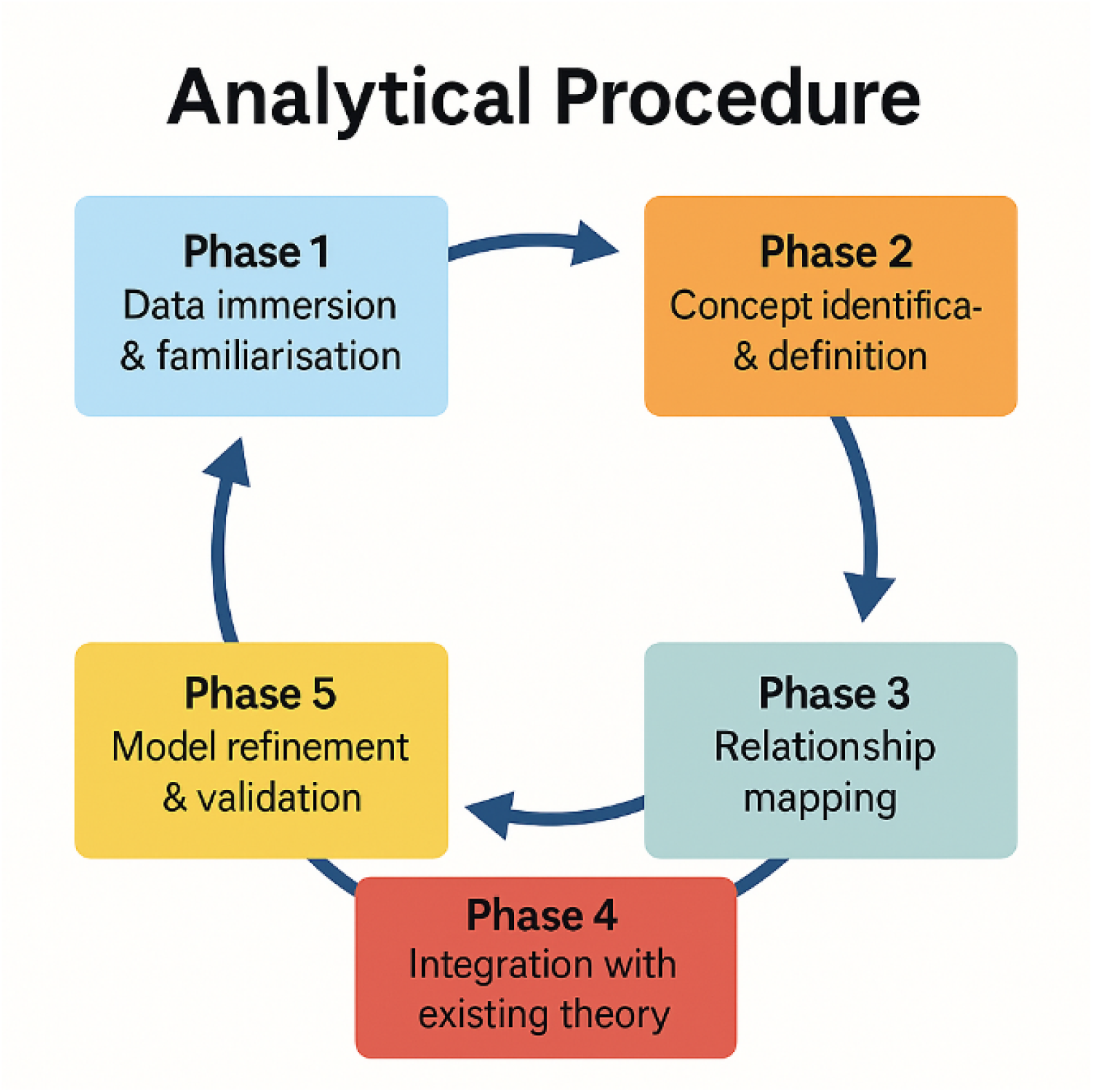
Analytical Procedure Showing the Five Phases of Model Development.

##### Phase 1: Data Immersion and Familiarisation

The lead author (LAN) systematically reviewed all primary studies including publications, transcripts, field notes, datasets, and protocols to identify recurrent patterns. Qualitative data were imported into NVivo 12 (QSR International) for systematic coding; quantitative datasets were managed using SPSS Version 28 (IBM Corp., Armonk, NY). All data sources were catalogued chronologically and by study design to facilitate temporal and methodological triangulation.

##### Phase 2: Concept Identification and Preliminary Framework Development

Core constructs were derived through iterative thematic coding. Initial open coding identified 87 discrete concepts, collapsed through axial coding into five higher-order constructs: (1) cultural and occupational context, (2) back pain beliefs, (3) coping strategies, (4) participant activation, and (5) self-management behaviours. Operational definitions were informed by the Back Beliefs Questionnaire (BBQ) [11,26], Fear-Avoidance Beliefs Questionnaire (FABQ) [30], and Patient Activation Measure (PAM) [28], and by culturally grounded insights from African settings [3,14,17,22,24,31,34,35]. A preliminary framework mapped hypothesised relationships between constructs based on temporal precedence (beliefs precede coping), theoretical logic (context shapes beliefs), and observed statistical associations (beliefs predict activation).

##### Phase 3: Systematic Indexing of Data against Emerging Constructs

All empirical data from the 17 contributing studies were systematically indexed against the five higher-order constructs. Each data segment was assigned to one or more construct categories. Qualitative data segments coded in NVivo 12 were cross-referenced with quantitative findings to ensure construct definitions were empirically grounded. Where constructs overlapped, for example, cultural norms directly shaping both beliefs and coping preferences construct boundaries were refined. Contradictory evidence was documented and examined for moderating conditions, producing a structured evidence map as the analytic foundation for Phase 4.

##### Phase 4: Integration with Existing Theoretical Frameworks

Emergent relationships were systematically compared with the Fear-Avoidance Model [30], Biopsychosocial Model [27], and Patient Activation Model [28]. This comparison identified convergence, pain-related fear predicts disability and critical divergences for LMIC contexts. The AC-BP-SMM positions context as the foundational driver rather than a background influence, and demonstrates that high activation fails to predict behaviour when structural barriers block enactment the knowledge-action gap. African-specific constructs (stoicism as virtue, spiritual attributions, occupational self-sacrifice, and collectivist responsibility) were explicitly incorporated.

##### Phase 5: Model Refinement through Internal Validation

The draft model underwent iterative refinement through internal consistency checking, testing against contradictory cases and outlier findings and preliminary member-checking with 23 Zambian nurses and physiotherapists through facilitated group discussions. Participants confirmed resonance with the knowledge-action gap (*“Yes! I teach patients to stay active but cannot do this myself because of no time”*). Feedback led to terminology refinement (*changing “compliance” to “behavioural enactment”*) and addition of economic stress as a moderator. These processes established internal validity, which informed the development of the Delphi instrument in Stage 2.

### 2.2 Stage 2: Content Validity Evaluation — the Delphi Technique

#### 2.2.1 Rationale for the Delphi Approach

The Delphi technique is a structured, multi-round expert consultation method designed to achieve consensus and evaluate the content validity of theoretical models and assessment instruments [38]. It is particularly appropriate for novel conceptual models where empirical validation is ongoing and expert judgement is required to confirm content representativeness, relevance, and clarity [39]. The Delphi approach was selected to formally evaluate whether the AC-BP-SMM’s constructs, relationships, and moderators adequately and comprehensively represent the domain of back pain self-management in African LMIC contexts. This study followed the Guidance on Conducting and REporting DElphi Studies (CREDES) recommendations [40], which provide methodological standards for Delphi design, conduct, and transparent reporting.

#### 2.2.2 Expert Panel Selection and Recruitment

A purposive sampling strategy was used to assemble an expert panel of 18 participants. Inclusion criteria required demonstrated expertise in at least one of the following domains: back pain assessment and management; musculoskeletal physiotherapy in LMIC settings; occupational health in sub-Saharan Africa; health behaviour theory and models; digital health intervention design; or healthcare policy in LMICs. Experts were recruited from 10 countries (Zambia, South Africa, Zimbabwe, Nigeria, Uganda, Ghana, United Kingdom, Canada, Australia, and the Netherlands) through professional networks, research databases (PubMed author searches), and snowball referrals. A minimum panel size of 10–15 experts is recommended for content validity studies; the panel comprised 18 experts (7 physiotherapists, 4 occupational health practitioners, 4 health behaviour researchers, and 3 policy/public health experts). All participants provided written informed consent.

#### 2.2.3 Delphi Instrument Development

Based on the draft AC-BP-SMM derived from Stage 1, a structured Delphi questionnaire was developed comprising 17 items organised across the five model components and four moderators. Each item described a specific construct, relationship, or model feature. Experts were asked to rate each item on three dimensions: (1) Relevance; the degree to which the item represents a necessary component of the model (4-point scale: 1 = not relevant, 2 = somewhat relevant, 3 = quite relevant, 4 = highly relevant); (2) Clarity; the degree to which the item statement is understandable and unambiguous (4-point scale: 1 = not clear, 4 = very clear); and (3) Representativeness; whether the item is representative of the construct as defined (yes/no with comment). Open-ended response fields enabled qualitative feedback on each item and on the model overall. The instrument was piloted with two academic physiotherapists not included in the main panel for clarity and face validity.

#### 2.2.4 Delphi Rounds and Data Analysis

The Delphi study was conducted over two rounds, administered electronically via REDCap (Research Electronic Data Capture, Vanderbilt University). Round 1 presented all items for initial rating with open feedback. Between rounds, quantitative results were summarised and qualitative feedback was thematically analysed to develop revised items. Participants received anonymised group statistics for their reference in subsequent rounds. The process continued until stability of responses was achieved or predefined consensus thresholds were met.

Content Validity Index (CVI) scores were calculated following established methodology [41,42]: the Item-level CVI (I-CVI) was calculated as the proportion of experts rating an item as “quite relevant” or “highly relevant” (scores 3 or 4) on the relevance scale. The Scale-level CVI (S-CVI/Ave) was calculated as the average of all I-CVI scores across items. Consensus was defined a priori as I-CVI ≥ 0.80 per item and S-CVI/Ave ≥ 0.90 for the overall model. Items failing to achieve consensus were revised or removed based on expert feedback. Fleiss’ kappa was calculated to assess inter-rater agreement beyond chance.

### 2.3 Analytical Outputs and Data Presentation

The synthesis generated quantitative and qualitative outputs. Quantitative findings were presented using prevalence estimates with 95% confidence intervals, knowledge-action gap calculations as difference scores with subgroup stratification, and odds ratios for moderator effects. Qualitative findings were presented using thematic typologies, representative participant quotations, and narrative descriptions of lived experiences. Delphi results were presented as I-CVI and S-CVI/Ave scores by round, with expert feedback themes summarised narratively. Visual models synthesised evidence through sequential pathway diagrams, moderator illustrations, and the integrated AC-BP-SMM diagram.

### 2.4 Quality and Rigour Considerations

The synthesis maintained rigour through methodological triangulation across qualitative, quantitative, and intervention studies; temporal depth spanning 15 years; sample diversity across healthcare roles and settings; theoretical transparency; and external corroboration through alignment with LMIC literature from Uganda [24,25], Nigeria [31,34], South Africa [17,22], Ghana, Kenya, Pakistan [35], and Brazil [32]. The Delphi stage enhanced content validity through systematic expert consensus evaluation across multiple rounds.

### 2.5 Ethical Considerations

All contributing empirical studies received ethical approval from relevant institutional review boards in Zambia and partner universities. This conceptual synthesis used only published aggregate findings and did not involve new data collection. The Delphi study received ethical approval from University of Zambia Health Sciences Research Ethics committee (UNZAHSREC), Protocol ID 2023271507. Expert panel participants provided written informed consent and were informed of their right to withdraw at any time. All responses were anonymised in analyses and reporting.

## 3. Results

### 3.1 Overview of the AC-BP-SMM

Fig 2 illustrates the sequential pathway from context to self-management outcomes, moderated by four key factors. The model positions cultural and occupational context as the foundational determinant shaping all subsequent components. Unlike Western models where context is a background influence, the AC-BP-SMM foregrounds it as the primary driver of beliefs, coping, activation, and behaviour.

**Fig 2.**
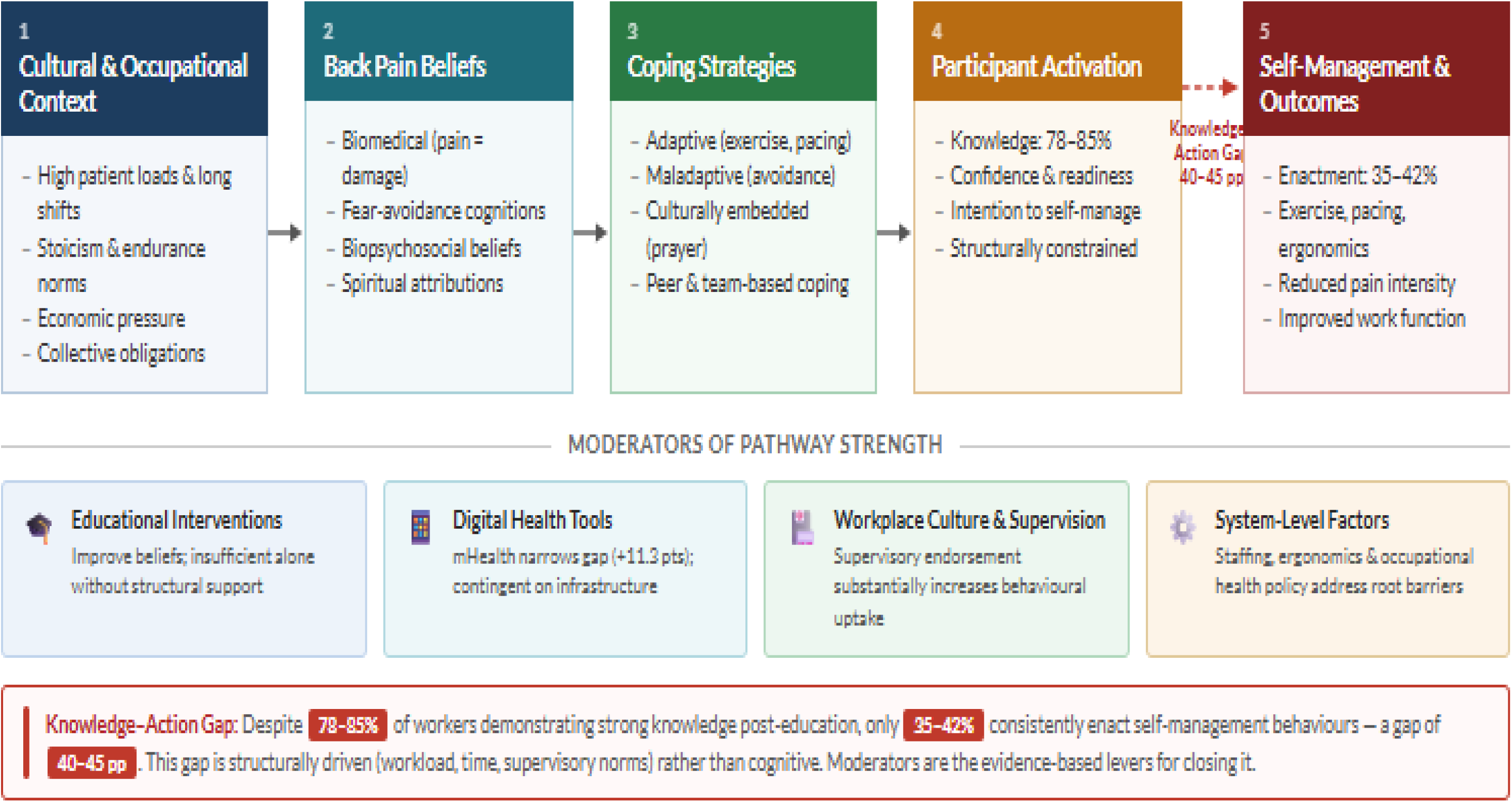
Sequential Pathway from Context to Self-Management Behaviours with Four Moderators Influencing Pathway Strength.

The model proceeds through five interconnected components: (1) Cultural and Occupational Context, encompassing workplace demands, cultural norms, economic pressures, and social expectations; (2) Back Pain Beliefs, ranging from biomedical to biopsychosocial, including fear-avoidance cognitions and spiritual attributions; (3) Coping Strategies, including adaptive, maladaptive, and culturally embedded approaches; (4) Participant Activation, conceptualised as knowledge, confidence, and readiness constrained by contextual realities; and (5) Self-Management Behaviours and Outcomes. Four moderators; educational interventions, digital health tools, workplace culture, and system-level factors influence pathway strength and the capacity to bridge the knowledge-action gap.

### 3.2 The Knowledge-Action Gap

Across multiple studies, a persistent and quantifiable discrepancy emerged between knowledge possession and behavioural enactment (Figure 3). Following educational interventions, 78–85% of healthcare workers demonstrated strong biomedical and biopsychosocial back pain knowledge evidenced by significant improvements in Back Beliefs Questionnaire scores (mean increase of 15–22 points on a 100-point scale; p<0.001) [5,26,33]. Yet only 35–42% of the same workers reported consistently implementing evidence-based self-management strategies over 3–6 month follow-up periods [6–8], creating a 40–45 percentage point gap. Among nurses who received back pain education (n=387 across three studies), 82% demonstrated strong knowledge at 3-month follow-up but only 38% reported consistent self-management behaviour, yielding a 44 percentage point gap [5–7,26,33]. In-depth interviews (n=87 across five studies) confirmed this gap was not due to forgetting or lack of motivation but to specific, concrete, and contextual barriers.

> *“I know I should do exercises every day. I even teach my patients this. But after a 12-hour shift with 60 patients, lifting, bending constantly, I just collapse at home. I have no energy left.” [Nurse, tertiary hospital, 8 years’ experience]*

**Fig 3.**
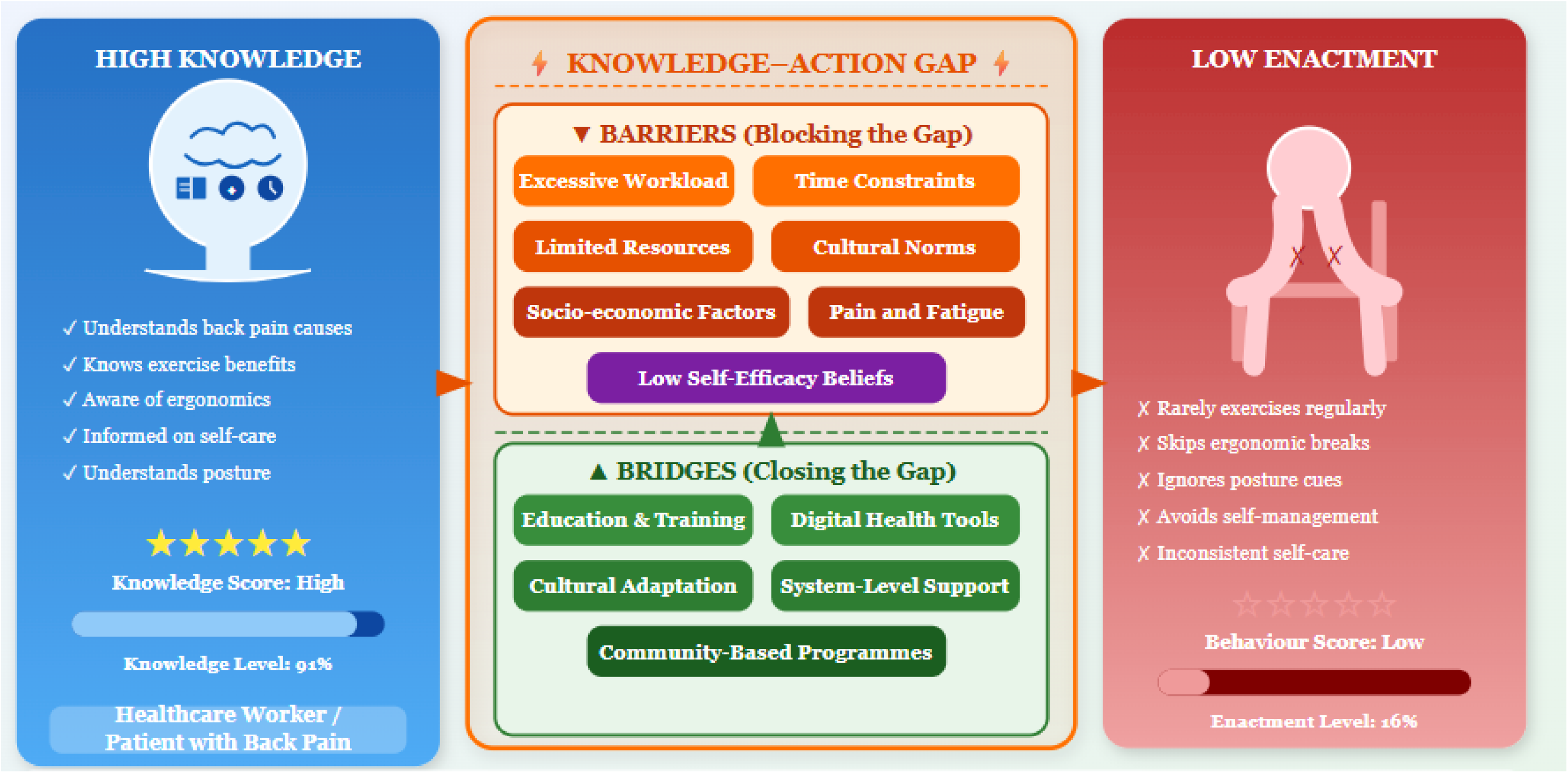
Knowledge-Action Gap in Back Pain Self-Management: High Knowledge vs. Low Enactment, with Barriers Widening the Gap and Bridges Proposed to close it.

> *“The physiotherapist showed me stretches. I remember them all. But when can I do them? Morning I’m preparing children for school, then shift starts, no break, then home to cook, clean, fetch water. Where is the time?” [Nursing assistant, rural health centre, 4 years’ experience]*

> *“My supervisor would see it as laziness if I took breaks to stretch. She says we’re here to work, not to pamper ourselves. Even when my back is screaming, I keep going because I need this job.” [Nurse, secondary hospital, 6 years’ experience]*

Table 2 demonstrates that the knowledge-action gap varies with contextual supports, not individual traits. It is structurally driven, reflecting systemic barriers rather than cognitive or motivational deficits.

**Table 2.** Knowledge-Action Gap Magnitude by Moderating Factor.

| Moderating Factor | Knowledge (%) | Behaviour (%) | Gap (pp) | Key Quotation | Mechanism |
| --- | --- | --- | --- | --- | --- |
| **Overall (Baseline)** | 78–85 | 35–42 | 40–45 | *"I know I should do exercises... but after a 12-hour shift with 60 patients I just collapse at home."* | Core gap across multiple sites; not due to knowledge deficit but contextual strain. |
| **Workload Effects** | 85 | 57 (<50 pts/shift) 25 (>50 pts/shift) | 28 (low) 58 (high) | *"After a 12-hour shift... no energy left."* | Excessive workload directly inhibits enactment; creates exhaustion and time scarcity. |
| **Supervisory Support** | 84 (support.) 81 (unsupport.) | 53 (support.) 29 (unsupport.) | 31 (support.) 52 (unsupport.) | *"My supervisor would see it as laziness if I took breaks to stretch."* | Supportive culture legitimises self-care; authoritarian supervision reinforces moral distress. |
| **Community/Peer Support** | 83 (with) 80 (without) | 48 (with) 32 (without) | 35 (with) 48 (without) | *"My colleague reminds me to stretch, and we cover for each other."* | Peer accountability and shared problem-solving enhance behavioural translation. |
| **Digital Tool Access** | 84 (with) 82 (without) | 55 (with) 35 (without) | 29 (with) 47 (without) | *"WhatsApp reminders help me remember to move... but during busy shifts I can't always follow."* | Digital reinforcement aids motivation but cannot overcome structural workload barriers. |
| **Economic Stress** | 80 (low) 83 (high) | 46 (low) 30 (high) | 34 (low) 53 (high) | *"Even when my back is screaming, I keep going because I need this job."* | Financial insecurity overrides health intentions, widening the gap. |
| **Cultural Norms & Fear** | — | — | — | *"I know rest makes it worse... but what if I damage something permanently?"* | Fear of injury and cultural endurance expectations discourage self-management. |

### 3.3 Component 1: Cultural and Occupational Context

Cultural and occupational context emerged as the foundational determinant shaping all subsequent pathway components. Four contextual dimensions were identified: workplace demands (high patient loads, long shifts, inadequate staffing, physically demanding tasks); cultural norms (stoicism as virtue, pain as inevitable, professional self-sacrifice, collective responsibility); economic pressures (low wages requiring multiple jobs, financial insecurity, limited sick leave); and social expectations (family caregiving responsibilities, gender role expectations, community status tied to employment) [11,13–19,22,24,31,33–35]. These were not merely background influences but active shapers of beliefs and behaviours. Cultural norms determined whether pain was viewed as requiring intervention or as something to be endured silently. Economic pressures created competing priorities where immediate income generation superseded long-term health preservation. This positioning of context as primary not a moderator of individual factors represents a key theoretical departure from Western models.

### 3.4 Component 2: Back Pain Beliefs

Back pain beliefs existed on a spectrum from biomedical to biopsychosocial, with many healthcare workers holding mixed or contradictory views. Biomedical beliefs perceptions that pain signified structural damage, required rest, and risked progression if activity continued were common [11,14,26] and reinforced by the physical reality of care work. Fear-avoidance beliefs were present among workers who believed movement could worsen tissue damage [11,26,30]; exposure to educational interventions and evidence-based messaging increased acceptance of biopsychosocial constructs [5,26,33]. A consistent finding was the coexistence of biomedical and biopsychosocial beliefs in the same individuals’ nurses might encourage patients to stay active while personally believing their own pain required rest reflecting the influence of occupational realities and cultural expectations on personal health behaviour. Beliefs strongly influenced coping styles but were not sufficient predictors of behaviour due to contextual constraints [25,32,35].

### 3.5 Component 3: Coping Strategies

Healthcare workers employed three broad categories of coping strategies that is adaptive, maladaptive, and culturally embedded as summarised in Table 3. Adaptive coping (maintaining activity, pacing, stretching) was most common among workers with biopsychosocial beliefs or exposure to back pain education [5,6,11,26], but consistently constrained by fatigue, long shifts, limited time, and inadequate workspace [14,15,17,22,24,31,34,35]. Maladaptive coping was frequently context-driven emerging from heavy workloads or limited autonomy rather than simply cognitive. Culturally embedded coping (prayer, spiritual surrender, quiet endurance) reduced emotional distress but could inadvertently reduce engagement in active self-management [3,14,24,31,32,35].

**Table 3.** Typology of Coping Strategies Used by Healthcare Workers Managing Back Pain in LMIC Settings.

| Coping Category | Description | Examples | Key Influences / Notes |
| --- | --- | --- | --- |
| **Adaptive Coping** | Behaviours that support functional recovery and align with evidence-based back pain management. | Maintaining activity within tolerance, stretching/exercise, pacing and activity modification, heat therapy, adjusting lifting techniques. | Promoted by biopsychosocial beliefs and education [5,6,11,26]; constrained by fatigue, long shifts, limited time, inadequate workspace [14,15]; similar barriers across SSA [17,22,24,31,34,35]. |
| **Maladaptive Coping** | Behaviours that restrict functional recovery or reinforce fear and inactivity. | Prolonged rest, activity avoidance, fear-based movement restriction, over-reliance on analgesics, medicalising routine symptoms. | Linked to biomedical/fear-avoidance beliefs [11,26,30]; often context-driven rather than solely cognitive; response to structural constraints and limited autonomy. |
| **Culturally Embedded Coping** | Coping shaped by cultural, spiritual, and social norms within African healthcare settings. | Prayer and spiritual surrender, normalising pain as professional duty, social/team-based coping, quiet endurance to avoid appearing weak. | Reduces emotional distress but may reduce active self-management [3,14,31]; documented across Uganda, Ghana, Pakistan, Brazil [24,32,35]; not reflected in Western coping frameworks. |

### 3.6 Component 4: Participant Activation

Participant activation knowledge, confidence, and readiness to self-manage 0pis highly context-dependent and structurally constrained in LMIC healthcare settings, unlike the individually determined construct conceptualised in high-income settings [28]. Three activation patterns consistently emerged: (1) high knowledge but low confidence; (2) high intention but no opportunity; and (3) moderate activation suppressed by workplace culture. Table 4 summarises these patterns and their predictors.

**Table 4.** Activation Patterns and Predictors among Healthcare Workers in LMIC Settings Part A. Common Activation Patterns.

| Pattern | Description | Key Drivers |
| --- | --- | --- |
| **High Knowledge, Low Confidence** | Workers possess strong understanding of evidence-based principles but lack confidence in applying them within real-world constraints. | Persistent pain despite correct self-management; limited workspace; absence of role models; fear of colleague judgement; prior dismissive healthcare experiences. |
| **High Intention, No Opportunity** | Strong motivation exists but structural barriers prevent behavioural execution. | Excessive workload and staffing shortages; fatigue from $\geq 12$-hour shifts; competing domestic duties; lack of breaks; financial limitations. |
| **Moderate Activation Suppressed by Culture** | Adequate knowledge and motivation undermined by workplace norms discouraging self-care. | Cultural expectation to prioritise patients over self; perceived impropriety of on-duty stretching; unsupportive supervision; stigmatisation of breaks; lack of occupational health policy. |

Part B. Predictors of Activation Level
| Predictor | Direction | Effect on Activation |
| --- | --- | --- |
| **Strong Biopsychosocial Beliefs** | ↑ | Understanding pain as multifactorial increases confidence and motivation ($\beta = 0.38$, $p < 0.001$). |
| **Adaptive Coping Strategies** | ↑ | Problem-focused coping linked to higher activation ($r = 0.42$, $p < 0.001$) by reinforcing self-efficacy. |
| **Workplace Support** | ↑ | Supervisory endorsement and peer normalisation of self-care predict higher activation, even under high workload. |
| **Digital Health Access** | ↑ | WhatsApp-based interventions raised activation by 11.3 points (95% CI 8.1–14.5); sustained 3-month effects. |
| **High Occupational Stress** | ↓ | Chronic stress depletes cognitive and emotional resources, weakening the belief–activation link ($\beta = -0.24$, $p < 0.01$). |
| **Heavy Workload** | ↓ | Patient-to-nurse ratios $> 1:20$ reduce activation by 15%, eroding capacity for self-management. |
| Low Job Autonomy | ↓ | Lack of control over pace, breaks, or task modification suppresses activation, particularly among junior staff. |

### 3.7 Component 5: Self-Management Behaviours and Outcomes

Self-management behaviours included exercise, pacing, stretching, hot/cold therapy, ergonomic changes, and self-care routines [13,14,17,22,24,26,31–35,43,44]. While workers used a range of strategies, frequency, consistency, and duration remained low constrained by contextual factors including fatigue, space limitations, and ward culture. Workers with higher activation and supportive environments reported reduced pain intensity, fewer severe episodes, lower interference with work, and greater functional engagement. Outcomes varied widely and were moderated by staffing adequacy and physiotherapy access (Fig 4).

**Fig 4.**
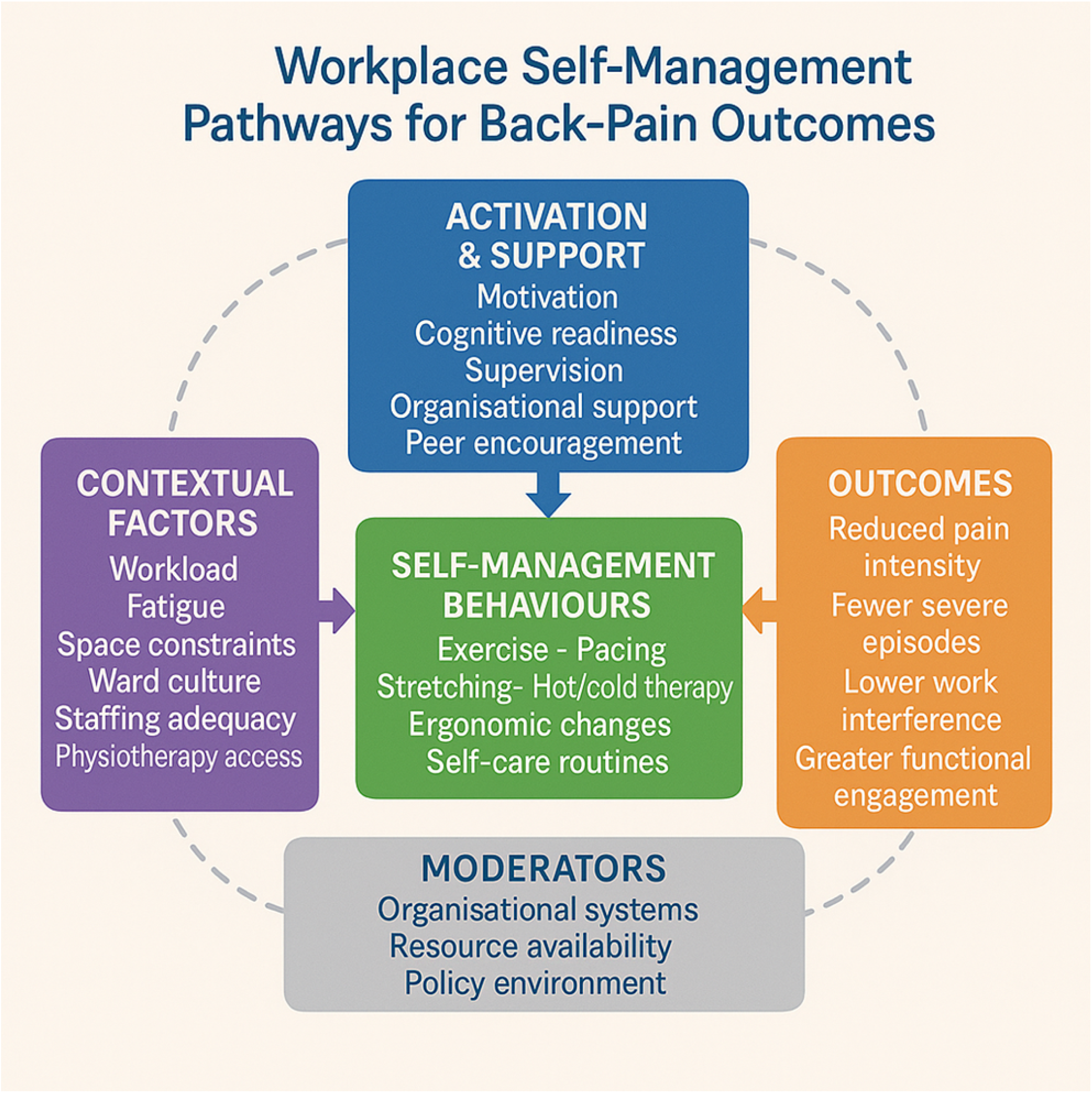
Healthcare Workers’ Self-Management Behavioural Pathways: Flow from Activation to Behaviour Enactment and Outcomes, with Contextual Factors as Barriers and Moderators as Enablers.

### 3.8 Moderators Influencing the Pathway

Four moderators determine whether healthcare workers successfully translate activation into behaviour (Fig 5). Educational interventions improve knowledge and shift beliefs but result in limited behavioural change without structural support [5,22,26,33]. Digital health tools, particularly WhatsApp-based programmes, enhance activation through flexible access, ongoing reinforcement, and peer encouragement [6,45–49] but cannot compensate for structural constraints during clinical shifts; evidence from LMIC hospital settings confirms that even well-designed digital platforms encounter uptake barriers rooted in workload and infrastructure [48,50]. Workplace culture and supervisory support determine whether activated workers feel permitted to engage in self-care; supportive supervision substantially increases behavioural uptake [17,22,51]. System-level reforms adequate staffing, ergonomic equipment, occupational health services address foundational barriers without which activation remains potential rather than practice [10,17,23,36,37]. Greatest impact occurs when all four moderators are activated simultaneously.

**Fig 5.**
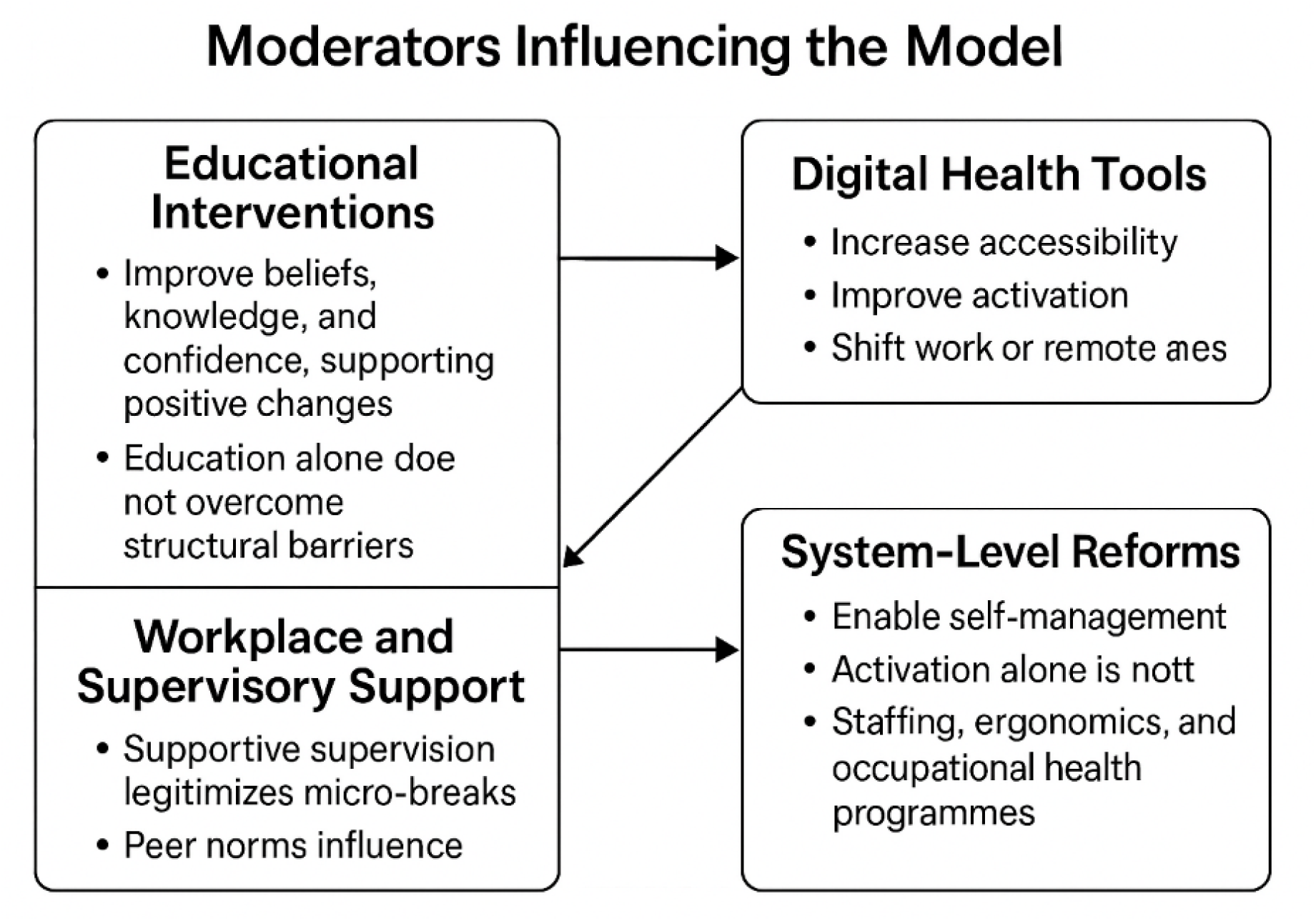
Moderators Influencing the LMIC Healthcare Worker Back Pain Pathway: Educational Interventions, Digital Health Tools, Workplace Culture, and System-Level Reforms as Enablers of Behaviour Change.

### 3.9 The Complete AC-BP-SMM

Fig 6 presents the complete integrated AC-BP-SMM. The model integrates all five components into a cohesive cyclical framework, with cultural and occupational context as the foundation, a sequential pathway from beliefs through coping and activation to self-management behaviours, and four moderators operating across the pathway. The knowledge-action gap is identified as the critical juncture between activation and behaviour. The model’s central contribution is demonstrating that this gap arises not from individual deficits but from structural and cultural constraints reframing the intervention problem from individual behaviour change to structural enablement.

**Fig 6.**
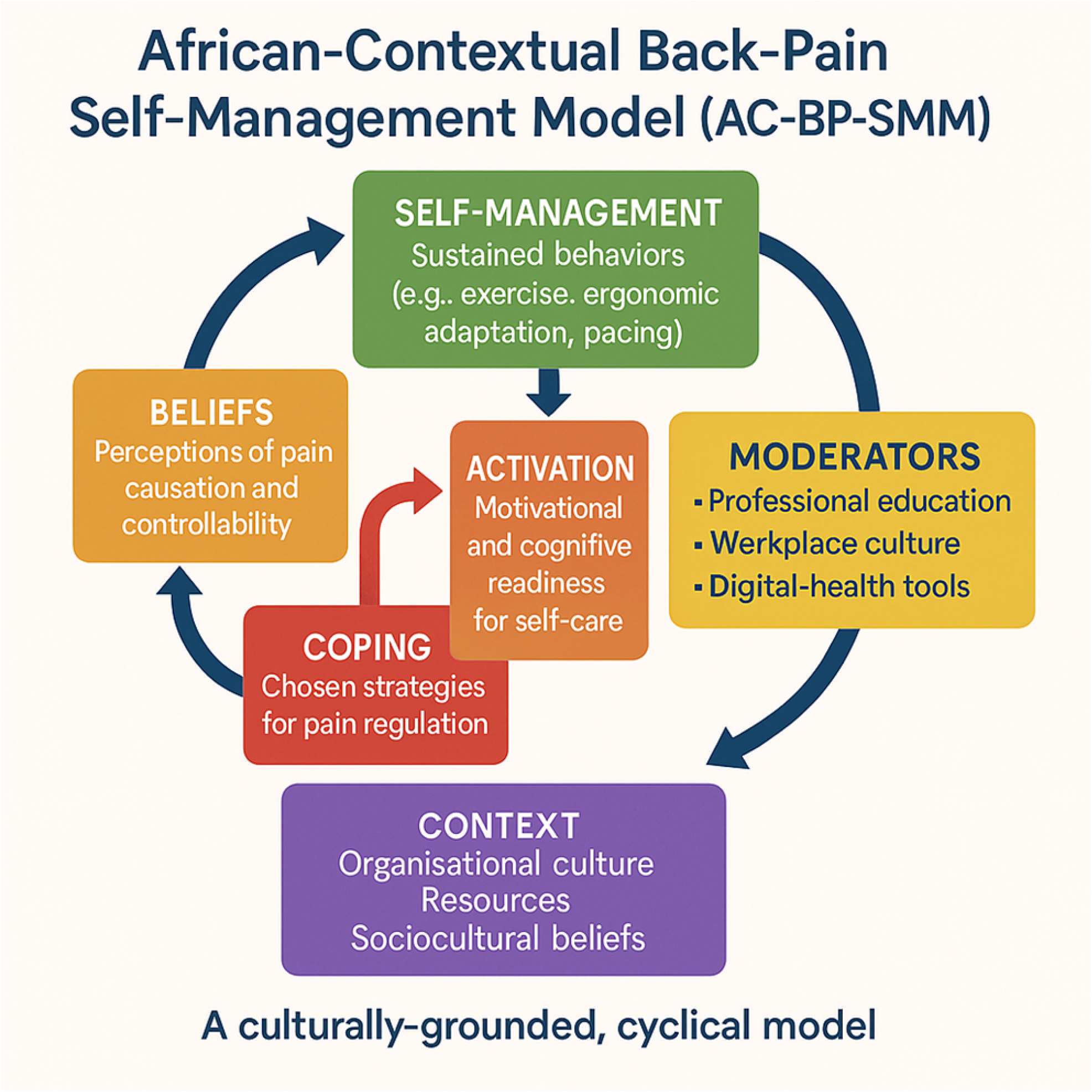
The Complete AC-BP-SMM: A Culturally-Grounded, Cyclical Model Showing All Five Components, Four Moderators, and the Knowledge-Action Gap.

### 3.10 Delphi Content Validity Results

The Delphi study was conducted over two rounds with 18 experts. Response rates were 100% (18/18) in Round 1 and 94.4% (17/18) in Round 2. In Round 1, 14 of 17 items achieved I-CVI ≥ 0.80. Expert feedback led to revision of three items primarily related to the operationalisation of the knowledge-action gap and the spiritual coping construct within the Back Pain Beliefs and Coping components. Following revision, Round 2 yielded I-CVI ≥ 0.80 for all 17 items. The overall S-CVI/Ave after the final round was 0.93 (17 items retained, 0 added, 0 removed). Inter-rater agreement was κ = 0.82 (95% CI 0.74–0.90), indicating substantial consensus. Table 5 summarises CVI scores by model component and item. Principal qualitative themes from expert feedback included: (i) strong endorsement of the knowledge-action gap construct as a critical and novel contribution; (ii) recommendations to more explicitly operationalise the spiritual coping construct to distinguish spiritual endurance from active spiritual coping; (iii) suggestions to clarify the boundary between the Activation and Self-Management Behaviour components; and (iv) recommendations to strengthen the articulation of digital health tools as a moderator rather than a component. These were incorporated in subsequent model revisions.

**Table 5.** Content Validity Index (CVI) Scores by Model Component and Item across Delphi Rounds.

| Model Component / Item | Item Description | Round 1 CVI | Round 2 CVI | Round 3 CVI | Retained |
| --- | --- | --- | --- | --- | --- |
| **Context Component** | Workplace demands (workload, staffing, and shift patterns) shape self-management capacity. | 0.89 | 0.94 | — | ✓ |
|  | Cultural norms (stoicism, spiritual coping, and endurance expectations) influence pain behaviour. | 0.83 | 0.89 | — | ✓ |
|  | Economic pressures and multiple job demands reduce capacity for self-management. | 0.86 | 0.92 | — | ✓ |
| **Back Pain Beliefs** | Biomedical beliefs (pain = damage) predict fear-avoidance and reduced activity. | 0.94 | 0.94 | — | ✓ |
|  | Biopsychosocial beliefs are associated with adaptive coping and higher activation. | 0.89 | 0.94 | — | ✓ |
|  | Spiritual attributions of pain are a culturally relevant determinant of self-management. | 0.72 | 0.89 | — | ✓ |
| **Coping Strategies** | Adaptive coping strategies are the most beneficial for long-term outcomes. | 0.94 | 0.94 | — | ✓ |
|  | Maladaptive coping is often structurally driven rather than purely cognitive. | 0.83 | 0.89 | — | ✓ |
|  | Culturally embedded coping (prayer, collective endurance) is a distinct LMIC construct. | 0.78 | 0.89 | — | ✓ |
| **Participant Activation** | Knowledge-action gap is the primary barrier between activation and behaviour. | 0.76 | 0.92 | — | ✓ |
|  | Workplace support and supervisory culture significantly moderate activation levels. | 0.89 | 0.94 | — | ✓ |
| **Self-Management Behaviours** | Exercise, pacing, and ergonomic modification are core self-management behaviours. | 0.94 | 0.94 | — | ✓ |
|  | Outcomes include pain intensity reduction, fewer episodes, and improved work function. | 0.89 | 0.94 | — | ✓ |
| **Moderators** | Educational interventions improve beliefs but require structural support for behaviour change. | 0.89 | 0.94 | — | ✓ |
|  | Digital health tools (mHealth/WhatsApp) narrow the knowledge-action gap. | 0.86 | 0.94 | — | ✓ |
|  | Workplace culture (supervisory support, peer norms) is a critical enabler of self-management. | 0.89 | 0.94 | — | ✓ |
|  | System-level factors (staffing, policy, and ergonomics) address foundational structural barriers. | 0.86 | 0.94 | — | ✓ |

## 4. Discussion

The AC-BP-SMM, developed through a rigorous 15-year framework synthesis and evaluated for content validity through the Delphi technique, represents a significant theoretical advancement in understanding back pain self-management in African healthcare settings. Three principal findings emerge. First, cultural and occupational context functions as a primary determinant, actively shaping beliefs, coping, activation, and behaviour at every stage of the pathway. Second, a quantifiable knowledge-action gap of 40–45 percentage points is the defining feature of back pain self-management in LMIC healthcare settings, not a knowledge deficit but a structural constraint gap. Third, the model achieved high content validity (S-CVI/Ave = 0.93), confirming that its constructs and relationships adequately and comprehensively represent the domain as judged by an international expert panel.

The AC-BP-SMM both builds upon and diverges from established frameworks. From the Fear-Avoidance Model [30], it retains the recognition that pain-related fear predicts disability, but extends this by showing that in LMIC contexts, avoidance often stems from structural constraints rather than catastrophic cognitions alone workers may avoid exercise not because they fear harm but because they lack time, space, or supervisory permission. This is consistent with evidence from Nigeria, Uganda, and Pakistan showing that fear-avoidant cognitions are modulated by resource scarcity and occupational powerlessness [24,31,35]. From the Biopsychosocial Model [27], it adopts integration of biological, psychological, and social factors while elevating cultural and social factors to primary determinants, a reorientation supported by qualitative evidence documenting how collectivist obligations and spiritual attributions of pain operate independently of psychological distress in African settings [3,14,15]. From the Patient Activation Model [28], it incorporates the activation construct but conceptualises it as structurally constrained rather than individually determined: high activation does not predict behaviour when external barriers block enactment, a finding corroborated by studies from South Africa, Ghana, and Brazil where high-knowledge workers with strong intentions consistently failed to enact self-management under adverse working conditions [17,22,32]. This distinction is critical: Western interventions aim to boost individual activation; LMIC interventions must simultaneously reduce structural barriers.

The formal content validity evaluation through the Delphi technique adds methodological rigour absent from previous LMIC back pain theoretical models. By demonstrating that an international expert panel with diverse professional backgrounds and regional expertise reached consensus (I-CVI ≥ 0.80 for all retained items), the model gains credibility as a content-valid framework suitable for operationalisation into assessment tools, intervention protocols, and evaluation frameworks. The expert-endorsed revisions, particularly the more explicit operationalisation of spiritual coping and digital health tools, strengthen the model’s utility for diverse LMIC populations.

The AC-BP-SMM’s unique contribution is its explicit focus on why knowledge does not translate to action in LMIC contexts and what moderators enable translation. This fills a gap in existing LMIC health behaviour theory, which often describes barriers without modelling the knowledge-action discrepancy or identifying actionable intervention targets. Abdullahi and Danazumi [31] have called for more African-led back pain research that foregrounds structural and cultural determinants; the AC-BP-SMM directly responds to this call by providing a validated theoretical architecture for such research. Compared to the Health Belief Model, the AC-BP-SMM moves beyond barrier identification to barrier mitigation strategies operationalised through specific moderators. Fatoye et al. [34] demonstrated that population-level incidence data alone cannot explain the persistence of back pain disability in constrained settings, reinforcing the need for contextualised explanatory models. Regarding digital health, the AC-BP-SMM’s positioning of technology as a moderator rather than a standalone solution aligns with emerging evidence that mHealth implementation in LMICs is contingent on organisational readiness and structural enablers [48–50], not merely device access. The hospital safety climate literature [51] similarly demonstrates that organisational culture, not individual knowledge, determines whether health workers adopt safe practices, directly paralleling the AC-BP-SMM’s finding that supervisory culture is a critical moderator of self-management enactment. Compared to task-shifting frameworks, it addresses what enables behaviour change in constrained contexts rather than simply who delivers care.

### 4.1 Implications

For intervention design, the model mandates multi-component approaches that simultaneously target beliefs (culturally adapted education) and barriers (digital tools, supervisory culture change, system reforms). Single-component educational approaches are insufficient and may paradoxically widen the knowledge-action gap by creating informed but structurally frustrated individuals. Micro-intervention exercises taking fewer than five minutes, buddy systems, and contextual cueing offer practical pathways within constrained LMIC environments. For occupational health policy, the model provides evidence that education alone is insufficient; policies must address workload, staffing adequacy, and workplace culture. Task-shifting back pain education to community health workers offers cost-effective expansion. Digital health strategies should prioritise offline-capable, SMS-based platforms accessible to low-literacy populations.

### 4.2 Limitations

Limitations include: the model’s grounding primarily in healthcare worker data from Zambia, requiring multi-country validation for cross-cultural applicability; correlational evidence for moderator effects requiring experimental manipulation for causal claims; partial reliance on self-report data potentially subject to social desirability bias; and the Delphi panel comprising experts from 10 countries, which, while diverse, may not fully represent the range of SSA contexts. Despite these limitations, the combined framework synthesis and Delphi approach offers substantial explanatory power and content validity evidence for the model.

### 4.3 Future Research Directions

The AC-BP-SMM requires empirical validation through: implementation trials testing whether targeting the knowledge-action gap outperforms education alone, using mixed methods and factorial designs for moderator testing; construct validity studies using structural equation modelling to test the sequential pathway relationships; cross-population validation in rural agricultural populations and urban informal workers; digital health research examining optimal modalities, content frequency, and low-literacy adaptations; and economic evaluations of model-guided interventions. Ongoing pilot research operationalising the AC-BP-SMM in rural Zambian communities through mHealth platforms, CHW support, and culturally adapted content will provide initial implementation evidence with findings expected in 2026.

## 5. Conclusion

The AC-BP-SMM is the first empirically grounded, content-valid conceptual model tailored for back pain self-management in African healthcare settings. Developed through rigorous framework synthesis of a 15-year research programme and evaluated through a multi-round Delphi technique, the model foregrounds cultural and structural context as primary determinants and operationalises the knowledge-action gap the persistent discrepancy between evidence-based knowledge and actual self-management behaviour as a measurable, structurally driven outcome. By achieving expert consensus (S-CVI/Ave = 0.93), the model has demonstrated the content validity necessary for operationalisation into assessment instruments, intervention protocols, and evaluation frameworks. The model’s practical mandate is clear: effective LMIC back pain interventions must simultaneously target beliefs and structural barriers through multi-component approaches. Education alone is insufficient and may paradoxically widen the gap it seeks to close. The AC-BP-SMM provides a theoretical foundation for the next generation of culturally appropriate, structurally sensitive interventions in Africa and beyond, applicable to healthcare workers, agricultural labourers, urban informal workers, and other LMIC populations experiencing the same fundamental tension between health knowledge and contextual constraints on action.

## Declarations

## Data Availability

All relevant data are within the manuscript and its Supporting Information files.

## Acknowledgements

The authors thank all healthcare workers who participated in the studies contributing to this synthesis. We acknowledge the University of Zambia and collaborating institutions for their institutional support. Special thanks to the nurses, physiotherapists, and caregivers in Zambia who generously shared their experiences and insights over the 15-year research programme, and to the expert panel members who participated in the Delphi evaluation.

## Ethics Approval and Consent to Participate

All contributing empirical studies received ethical approval from relevant institutional review boards in Zambia and partner universities. The Delphi study received ethical approval from the University of Zambia Health Sciences Research Ethics Committee (UNZAHSREC), Protocol ID 2023271507. All expert panel participants provided written informed consent.

## Consent for Publication

Not applicable.

## Availability of Data and Materials

The data supporting the framework synthesis are derived from published studies listed in the reference list. De-identified aggregate data and synthesis materials are available from the corresponding author (LAN) upon reasonable request. Delphi study data (anonymised) are available from the corresponding author upon reasonable request.

## Author Contributions

LAN: Conceptualisation, Data curation, Formal analysis, Investigation, Methodology, Project administration, Supervision, Validation, Visualisation, Writing — original draft, Writing — review & editing. GM: Conceptualisation, Investigation, Writing — original draft, Writing — review & editing. OJB: Conceptualisation, Writing — original draft, Writing — review & editing. MMM: Conceptualisation, Investigation, Supervision, Writing — original draft, Writing — review & editing. All authors have read and approved the final version of the manuscript.

## Abbreviations

AC-BP-SMM: African-Contextual Back Pain Self-Management Model
BBQ: Back Beliefs Questionnaire
CHW: Community Health Worker
CI: Confidence Interval
CVI: Content Validity Index
FABQ: Fear-Avoidance Beliefs Questionnaire
I-CVI: Item-level Content Validity Index
LMIC: Low- and Middle-Income Country
mHealth: Mobile Health
NVivo: Qualitative Data Analysis Software (QSR International)
OR: Odds Ratio
PAM: Patient Activation Measure
S-CVI/Ave: Scale-level Content Validity Index (Average)
SPSS: Statistical Package for the Social Sciences (IBM Corp.)
SSA: Sub-Saharan Africa
WHO: World Health Organisation.

## Supplementary Materials

Supplementary materials are available from the corresponding author (Loveness A. Nkhata) upon reasonable request: (i) detailed coding framework from Phase 2 of the framework synthesis; (ii) evidence mapping matrix from Phase 3; (iii) member-checking discussion guide from Phase 5; (iv) Delphi study instrument (all rounds); (v) round-by-round Delphi response data and expert feedback themes.

